# Are fast test results preferable to high test sensitivity in contact-tracing strategies?

**DOI:** 10.1101/2021.02.17.21251921

**Authors:** Jonas L. Juul, Kaare Græsbøll

## Abstract

Across the world, countries are fighting to reduce the spread of COVID-19. The backbone of the response is a test-trace-isolate strategy, where suspected infected get tested and isolated and possible secondary cases get traced, tested and isolated. Because more accurate tests often take longer to analyze, and the benefits of contact tracing are strengthened by rapid diagnosis, there exists a trade-off in test sensitivity and test waiting time in test-trace-isolate strategies. Here we ask: How many false negatives can be tolerated in a rapid test so that it reduces transmission better than a slower, more accurate test? How does this change with contact tracing efficiency and test waiting time? We find that a rapid, less sensitive test performs best for many test-parameter choices and that this is true even for modest contact tracing efficiency. For COVID-19-like viral parameters, a test with 40% false negatives and immediate result might reduce transmission as well as a test with no false negatives and a 3-day waiting time. Our analysis suggests employing rapid tests to reduce test waiting times as a viable strategy to reduce transmission when testing infrastructure is under stress.

## I. INTRODUCTION

Rapid diagnosis and isolation of COVID-19 cases is critical in reducing further transmission of the virus. Since a large fraction of infections take place before the infected develops symptoms,^1–3^ isolation following the onset of symptoms is not sufficient to control the pandemic.^4,5^ To ensure rapid diagnosis and reduce transmission before symptom onset, the World Health Organization therefore recommends tracing, testing and isolating close contacts of COVID-19 positives.^6^

The success of the test-trace-isolate strategy depends on many factors. In addition to social factors such as the population being informed and complying with guidelines, successful rapid diagnosis and isolation depends heavily on the following 3 factors. *i*. Test waiting time: How long it takes from a person wants to get tested to the result of the test arrives and contact tracing begins. *ii*. Test sensitivity: How often a test returns a false negative. *iii*. Tracing efficiency: The fraction of secondary cases that are successfully found through contact tracing confirmed positives.

These 3 strategy parameters – test waiting time, test sensitivity, and tracing efficiency – are not necessarily fixed. A surge in cases might cause an increase in test demands. Higher test demand could in turn cause longer turnaround times as seen in some countries during the fall and winter of 2020.^7,8^ Longer turnaround times lead to slower tracing of secondary infections and thus a vicious cycle begins. The 3 strategy parameters are also not independent. Faster test results often come at the cost of lower sensitivity. PCR tests are highly accurate, but take significant time to analyze. Faster results can be achieved by pooling tests^9–11^ or using rapid antigen tests,^12^ lateral flow devices^13^ or saliva tests.^14^

Recent analyses concluded that test sensitivity is secondary to frequency and turnaround times in population-screening strategies^15,16^ as implemented by Slovakia in November 2020.^17^ In context of the more-widely applied test-trace-isolate strategies, rapid tests increase the effect of contact tracing. This raises the natural question: When is it better to get a fast result than an accurate result? How does this change with the efficiency of contact tracing efforts, and with turnaround times? Here we confront these questions.

## II. MODEL DESCRIPTION

To quantify the impact of test-trace-isolate strategies on growing epidemics, we simulate the branching structure of the chains of infections, also referred to as the “epidemic tree”.^18,19^ Our model lets us keep track of who infected whom, which is essential when simulating contact tracing and quarantining infectious people. In the model (Figure 1A), infected individuals give rise to new cases, unless quarantined following testing and tracing efforts.

**FIG. 1.**
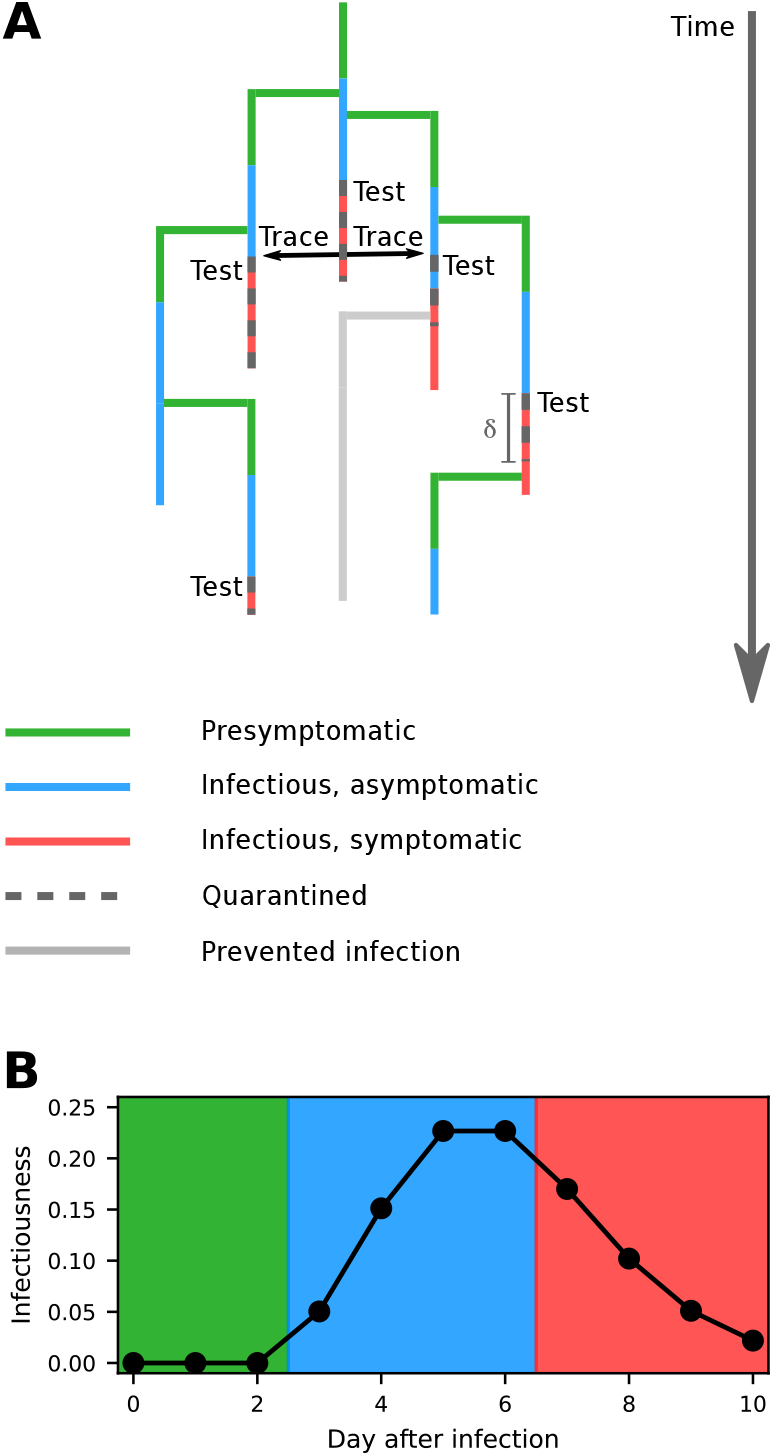
**A** Illustration of our model. Infectious people transmit the disease unless they are placed in quarantine following testing and tracing efforts. The success of these efforts are contingent on the three key parameters of the model: the test waiting time *δ*, test sensitivity 1 *− p*_false_, and tracing efficiency *p*_trace_. **B** Infectiousness over time, *P*_time_(*t*), in our model. Background colors specify whether the infected experiences symptoms and is infectious at the given time (see legend in Figure 1A) The value on the vertical axis for day *x* after infection is the probability that a given secondary case gets infected on day *x*. For asymptomatic cases, *P*_time_(*t*) is as depicted – the only difference being that the final, symptomatic, phase is replaced with an asymptomatic phase.

We initiate a simulation with some number of newly infected people, *N*_seed_ *∈* N. The simulation progresses in discrete timesteps, corresponding to days, and we make the simplifying assumption that every infected person, goes through the same phases before recovering: 3 days of being presymptomatic and noninfectious followed by 8 infectious days. As for COVID-19,^20^ some fraction of cases, *p*_asymp_, remain asymptomatic for the entire infectious period; all other infected cases experience symptoms starting on day 7 after infection.

In the absence of testing, tracing and isolation, an infectious person would give rise to *k* secondary cases. For each infected, we assume that *k* is drawn from the probability distribution *P* (*k*). When each of these *k* secondary cases is infected, we determine the time of infection by drawing an integer from the probability distribution *P*_time_(*t*). *P*_time_(*t*) takes positive values on the days where the infected is infectious and, mimicking COVID-19,^1,15^ peaks around symptom onset (Figure 1B). These infections take place unless the infected is in quarantine at the time the infection would occur.

Infectious people quarantine only when waiting for a test to be taken, receiving a test result, or after testing positive. In our model, an infectious person orders a test if either of two things happens: 1) The person is traced; 2) The person develops symptoms. In either case, the person orders a test immediately and then waits *δ* = *δ*_test_ +*δ*_result_ days for the result. The test waiting time is divided into *δ*_test_ days waiting for the test to be taken followed by *δ*_result_ days to receive the result. The test correctly identifies the case with probability equal to its sensitivity, 1 *− p*_false_, where *p*_false_ is the false negative rate. If the test comes back positive, each of the person’s secondary cases is traced with independent probability *p*_trace_. We obtain indistinguishable results when simulating the same *δ* with varying values of *δ*_test_ and *δ*_result_. Thus, the key parameters of the model are the test waiting time *δ*, test sensitivity 1 *− p*_false_, and tracing efficiency *p*_trace_.

Following *t*_max_ timesteps, we count the number of nodes that completed their whole infectious period, *n*_parents_. We also count the number of people these nodes infected, *n*_children_. The output of the simulation is the effective reproduction number of simulated disease: *R*_eff_ = *n*_children_*/n*_parents_.

## III. RESULTS

We use the model to examine the trade-off between test waiting time and test accuracy depending on the tracing efficiency. In our simulations, we therefore vary the parameters *δ*, 1 *− p*_false_, and *p*_trace_ and fix all other parameters (for *δ ≥* 1, we set *δ*_result_ = 1). For *P* (*k*) we choose a Poisson distribution with mean *R*_0_ = 2 (slightly lower than estimated in early stages of the pandemic^21,22^) and for *P*_time_(*t*) we choose the right-skewed distribution depicted in Fig. 1B. Finally, we choose *t*_max_ = 50.

To develop some intuition, let us first introduce the results we obtain when fixing the tracing efficiency, *p*_trace_ = 0.80. This constraint leaves 2 free parameters: The test sensitivity, 1 *− p*_false_, and the test delay, *δ*. We now compare the effective reproduction number 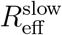 obtained by using a slow, but accurate test (parameters: 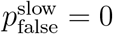 and some *δ*^slow^ *≥* 1 day) to the reproduction number, 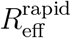, obtained with a less accurate, but rapid test (parameters: some 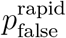 and *δ*^rapid^ = 0 days).

To evaluate whether speed or accuracy is to be preferred, we compute the difference in obtained effective reproduction numbers of the virus under the different choices of tests, 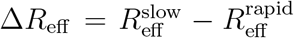. Let us choose some test delay, e.g. *δ*^slow^ = 2 days (perhaps corresponding to a PCR test with a 1-day waiting time to get tested and a subsequent 1-day waiting time to get the result). In this case, how will Δ*R*_eff_ depend on the risk of getting a false negative test result? For very high sensitivity (low *p*_false_), this faster test will be almost as accurate as the slower test it is being compared to. For this reason, the fast test will be preferable to the slower one (Fig. 2A top colorbar). If we now imagine slowly decreasing the test sensitivity, Δ*R*_eff_ will gradually increase until it reaches a breaking point where the slow and rapid tests reduce the effective reproduction number equally well: Δ*R*_eff_ = 0. Decreasing the sensitivity even further makes Δ*R*_eff_ positive, meaning that for this high probabilities of false negatives, the accurate test is to be preferred.

**FIG. 2.**
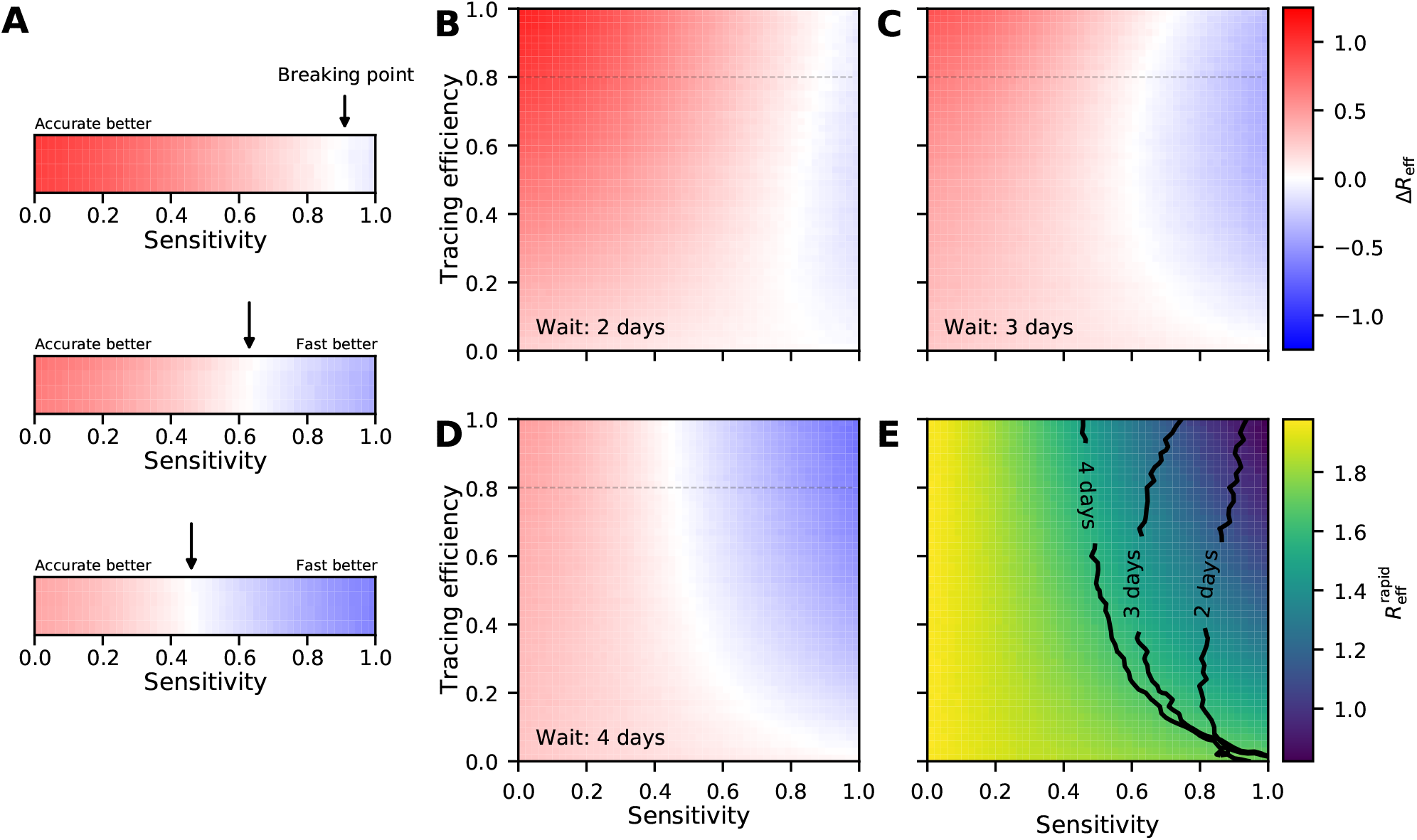
**A** Comparison of effective reproduction number, 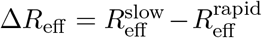, when using a slow but accurate test with sensitivity 1 (no false negatives), and a test waiting time of *δ*^slow^ days and using a rapid but less accurate test with a given sensitivity and test waiting time of *δ*^rapid^ = 0 days. In this inset, we assume that 80% of secondary cases are successfully traced following a positive test. The colorbars show results obtained for different choices of *δ*^slow^ (top colorbar: *δ*^slow^ = 2 days, middle: *δ*^slow^ = 3 days, bottom: *δ*^slow^ = 4 days), and the rapid-test sensitivity (horizontal axis in each colorbar). In each colorbar a breaking point separates sensitivity values favoring the slower, accurate test and values favoring the rapid test. **B** Comparison of effective reproduction numbers when the result of the slower test arrives after *δ*^slow^ = 2 days, as a function of the sensitivity and tracing efficiency. **C** Same as in B but with *δ*^slow^ = 3 days. **D** Same as in B, C but with the slower result arriving after *δ*^slow^ = 4 days. **E** Heatmap of the effective reproduction number obtained using the rapid, less accurate test, 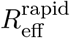. The breaking-point lines of B, C, D are plotted in black.

If we now make the same plot but with a higher waiting time to receive the slow test, the breaking point will move to a lower test sensitivity. The second colorbar in Fig. 2A shows the result for *δ*^slow^ = 3 days. The breaking point moved from sensitivity *≈* 0.91 to *≈* 0.63 with this single day increase in waiting time – corresponding to a fourfold increase in *p*_false_! Increasing the waiting time once more, setting *δ*^slow^ = 4, moves the breaking point further to the left. In this latter case, a 54% chance of a false negative is better than a 4 day wait for accuracy.

Having established some intuition for the simulations, we proceed to varying the third parameter: the tracing efficiency. By varying the tracing efficiency, for each choice of *δ*^slow^ we get 2-dimensional heatmaps instead of the one-dimensional colorbars presented in the previous paragraphs (Fig. 2B-D). In these heatmaps, the breaking points become white curves. Every point to the right of the breaking-point curve is a parameter combination where a faster test is preferable. Every point to the left of the breaking-point curve is a parameter combination that favors an accurate test. Notice how all the breaking-point curves start in the lower-right corner (where tests are completely accurate but no contact tracing is done) and how quickly they move to the left with increasing tracing efficiency. Figure 2E plots the obtained 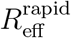. For each simulated choice of parameters, each computed *R*_eff_ is averaged over 10 simulations. For clarity, the heatmaps in Fig. 2 have been smoothed with a Gaussian filter.

## IV. DISCUSSION

Testing, tracing and isolating positive cases is central in many countries’ strategy to fight the current COVID-19 pandemic.^23–25^ We have demonstrated that there is a sizeable trade-off between test sensitivity and test waiting times in such strategies, and that it is often beneficial to prioritize test speed over test sensitivity. Moreover, we find that this benefit of rapid tests increases quickly with increases in test waiting times, and that even modest tracing efficiency unlocks the advantages of rapid tests. This indicates that additional waiting time for test results must be avoided and that it often makes sense to reduce test sensitivity in order to do so. It is to be expected that testing systems will occasionally get under stress during a pandemic, and having a way to avoid build-up of waiting times in this scenario is crucial. Designing such stress-relieve strategies presents an interesting direction for future research.

Some of the assumptions we have made can be questioned. Three such assumptions are: That quarantine hinters any transmission; that symptomatic individuals do not quarantine after testing (false) negative; and that the probability of getting a false negative result does not depend on the infected’s infectiousness at the time the test was taken. We note that all of these assumptions will favor reducing transmission by slower, more accurate tests: That people do not break isolation benefits the tests with long waiting times; false negative results leading to completely normal behavior damages only the tests that allow for false negative results; lastly, making false negatives less likely at high viral load would make the rapid tests more reliable early in the course of disease, when many new secondary cases could be avoided following diagnosis. That we have chosen our assumptions as to disfavor rapid low-sensitivity tests means that our results can be interpreted as conservative estimates of the benefits of reducing test waiting time with less sensitive tests.

Our choices of the probability distributions *P* (*k*) and *P*_time_(*t*) can also be questioned. A better choice for *P* (*k*) might be a heavy-tailed distribution that could account for superspreading behavior.^26,27^ Choosing a geometric distribution for *P* (*k*) with the same mean yields indistinguishable results (not shown). On the other hand, the choice of *P*_time_(*t*) influences results: Higher infectivity early in the course of disease makes the benefit of rapid tests greater (simulations not shown).

Two other limitations arise not from parameter choices, but the model itself. First, our model does not take depletion of susceptibles into account. Such effects are unimportant to what we were interested in: the effect of testing and tracing on the effective reproduction number at a given stage of the pandemic. Secondly, our implementation of contact tracing allows only for descendants of the confirmed positive to be traced. Not including two-way contact tracing or the possibility of identifying individuals through non-parents could cause us to underestimate the effect of contact tracing.^28–31^ This, and not taking into account possible beneficial effects on contact tracing that might be caused by infected being present at the test site when receiving the diagnosis, again biases our results against the rapid tests that so rely on tracing efforts.

Overall, our analysis suggests employing rapid tests to reduce test waiting times as a viable strategy to reduce transmission even at modest waiting times and contact tracing efficiency.

## Data Availability

The code and data necessary to reproduce the content of this paper is available on www.github.org/jonassjuul/TestTraceTradeoff

## ACKNOWLEDGMENTS

We thank SSI for contributing to funding this research. JLJ thanks Steven H. Strogatz for helpful discussions and Gorm G. Jensen for comments on an early version of the manuscript.

## AUTHOR CONTRIBUTIONS

KG and JLJ designed the research. JLJ designed the model, carried out the simulations, and wrote first draft of the manuscript. KG contributed to designing the model and made significant contributions to writing the manuscript.

## COMPETING INTERESTS STATEMENT

The authors declare that they have no competing interests.

## Notes

### Competing Interest Statement

The authors have declared no competing interest.

### Funding Statement

This work was partially funded by Statens Serum Institut (SSI).

